# Nasopharyngeal Carcinoma and Head and Neck Cancer in Patients with Type-2 Diabetes Mellitus receiving SGLT2I, DPP4I or GLP1a: A population-based cohort study

**DOI:** 10.1101/2024.01.14.24301285

**Authors:** Lifang Li, Oscar Hou In Chou, Kar Kei Mak, Yifan Yang, Cheuk To Chung, Guoliang Li, Catherine Chan, Wing Tak Wong, Tong Liu, Bernard Man Yung Cheung, Gary Tse, Jiandong Zhou

## Abstract

**Introduction:** Nasopharyngeal carcinoma (NPC) remains endemic in Asian regions, which risk factors were distinct from other head and neck (H&N) cancers. Anti-diabetic drugs has been proposed to reduce the risk of NPC. The associations between sodium glucose cotransporter 2 inhibitors (SGLT2I) versus dipeptidyl peptidase-4 inhibitors (DPP4I) and the risks of NPC and H&N cancer amongst type-2 diabetes mellitus (T2DM) patients remains unknown.

**Method:** This was a retrospective population-based cohort study including T2DM patients treated with either SGLT2I or DPP4I between 1^st^ January 2015 and 31^st^ December 2019 in Hong Kong. The primary outcome was new-onset NPC and other H&N cancer. The secondary outcome was cancer-related mortality. Propensity score matching (1:1 ratio) was performed using the nearest neighbour search. Multivariable Cox regression was applied to identify significant predictors.

**Results:** This cohort included 75,884 patients with T2DM, amongst whom 28,778 patients were on SGLT2I and 47,106 patients were on DPP4I. After matching (57556 patients), 106 patients developed NPC (0.18%), and 50 patients developed H&N cancer (0.08%). Compared to DPP4I, SGLT2I was associated with significantly lower risks of NPC (Hazard ratio [HR]: 0.41; 95% Confidence interval [CI]: 0.21-0.81) but not H&N cancer (HR: 1.00; 95% CI: 0.26-3.92) after adjustments. The result remained significant regardless of demographics and obesity. The association remained consistent in different risk models, matching approaches, and the sensitivity analysis.

**Conclusion:** This study provides real-world evidence that SGLT2I was associated with lower risks of NPC, but not H&N cancer compared to DPP4I after adjustments amongst T2DM patients.

## Introduction

Nasopharyngeal carcinoma (NPC) remains as major public health challenge due to their prevalence and mortality rate, especially in many Asian countries, with a reported incidence up to 50 per 100,000 population.^1^ This is in contrast to the prevalence of NPC in the Western countries which has a prevalence of less than 1 per 100. In general, head and neck (H&N) cancer are tumours derived from the mucosa of the aerodigestive tract from the nasal cavity to the larynx.^2^ One of the most important H&N cancer is NPC. NPC has a distinct epidemiology, aetiology and pathological characteristics compared to other H&N cancer.^3^ NPC is endemic in the southern region of China, as well as other parts of east and southeast Asia.^4^ Epstein-Barr virus (EBV) infection remained as the centre of the pathogenesis of NPC. Lifestyle and diet habits such as consuming preserved food which is more likely to be practised by Chinese population and smoking may become another potential risk factors for NPC.^5^ Since most patients with NPC are diagnosed with late-stage disease, and that patients diagnosed with metastatic NPC have poor prognosis, there remains to be a need for preventing the risks of new-onset NPC.^6^

Anti-diabetic drugs has been described in reducing the risk of NPC. In a Taiwan study, metformin was associated with a significantly lower risks of NPC compared to metformin non-user amongst T2DM patients.^7^ Sodium glucose cotransporter 2 inhibitors (SGLT2I) has demonstrated extensive anti-cancer effects across various malignancy, such that SGLT2I was associated with a reduced risks of cancer compared to placebo or other anti-diabetic drugs.^8–10^ EBV-encoded oncoprotein latent membrane protein 1 (LMP1) may increase the level of glucose transporter 1 and therefore enhance glucose uptake by the NPC cells and drives the Warburg effect.^6, 11^ As such, SGLT2I might have help prevent NPC by playing its role in the metabolic reprogramming. However, the effects of SGLT2I on NPC on a population scale remained uncertain. However, the association between other second-line anti-diabetic drugs such as SGLT2I and) dipeptidyl peptidase-4 inhibitors (DPP4I) remained scarce.

Since NPC remains endemic in Hong Kong, using the territory’s electronic health record data allows studying the association between SGLT2I and NPC using a data with large sample size, which is unlikely to be completed using a prospective study or randomised controlled trials.^12^ Hence, this study aimed to investigate the role of SGLT2I and DPP4I with new-onset NPC and H&N cancer in a cohort of T2DM patients from a territory-wide population cohort study.

## Methods

This study was approved by the Institutional Review Board of the University of Hong Kong/Hospital Authority Hong Kong West Cluster (HKU/HA HKWC IRB) (UW-20-250) and complied with the Declaration of Helsinki.

### Study design and population

This was a retrospective, territory-wide cohort study of T2DM patients who received their treatment with SGLT2I or DPP4I between 1st January 2015, and 31st December 2019, in Hong Kong. The follow-up period is continued until 31^st^ December 2020, or until the patients’ demise. A sensitivity analysis that included a cohort of patients on glucagon-like peptide-1 receptor agonist (GLP1a) between 1st January 2015 and 31st December 2019 was conducted to elucidate the relative effects among the second-line oral antidiabetic medications. The patients were identified from the Clinical Data Analysis and Reporting System (CDARS), a territory-wide database that centralizes patient information from individual local hospitals to establish comprehensive medical data, including clinical characteristics, disease diagnosis, laboratory results, and drug treatment details. The system has been used by local teams in Hong Kong to conduct comparative studies.^13–15^ The exclusion criteria of the study involved patients: 1) With prior NPC or H&N cancer 2) Without complete demographics 3) Under 18 years old **(Figure 1)**. This study employed an as-treat approach, including the censoring of patients at the discontinuation of treatment or switching between the comparison medications.

**Figure 1.**
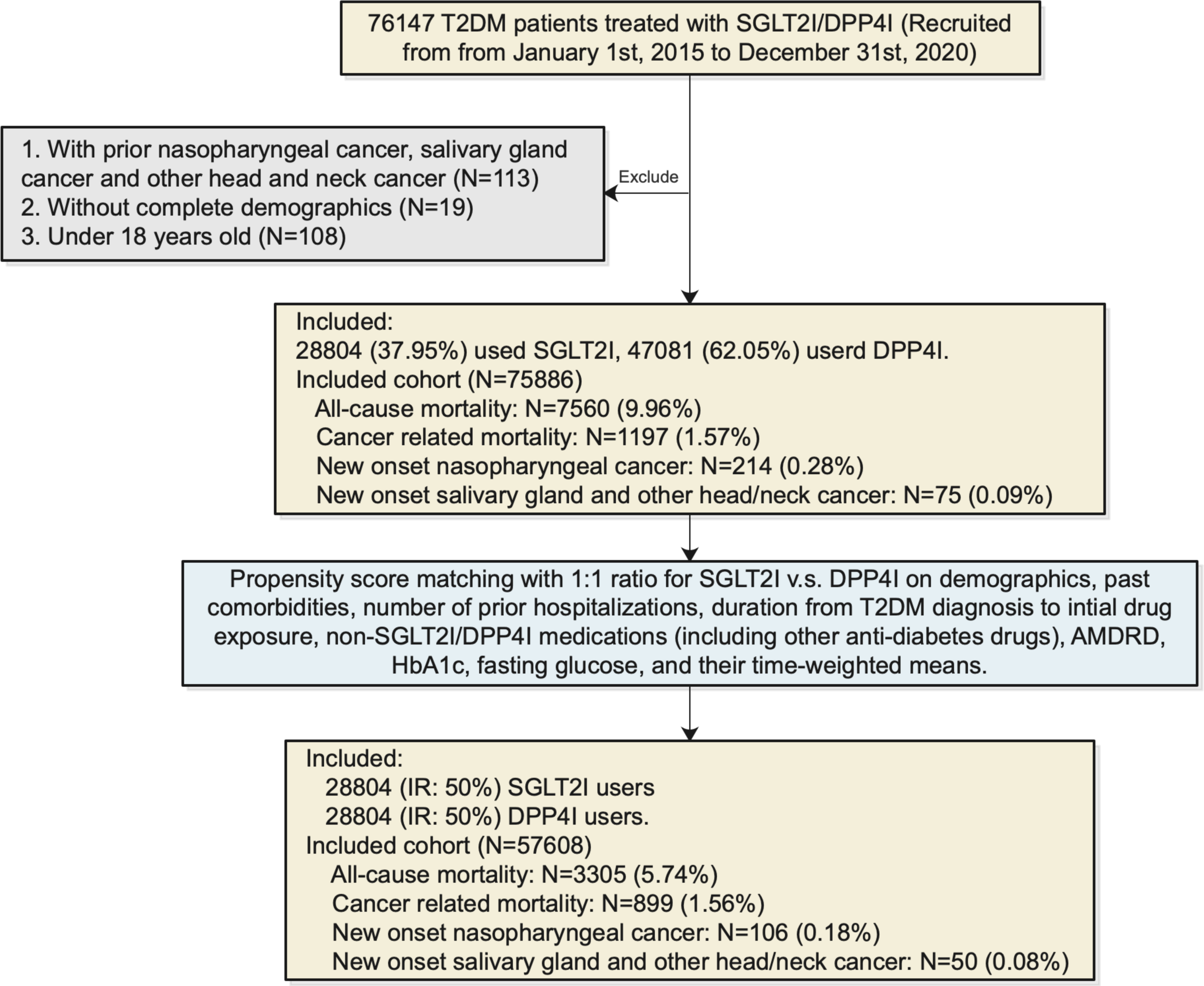
Procedures of data processing. SGLT2I: Sodium-glucose cotransporter-2 inhibitors; DPP4I: Dipeptidyl peptidase-4 inhibitors; AMDRD: abbreviated modification of diet in renal disease.

The demographic details of the patients, including gender and age at the initiation of drug use (baseline), clinical diagnoses and laboratory results as well as medications were extracted for this study. International Classification of Diseases Ninth Edition (ICD-9) codes (**Supplementary Table 1**) were used to identify the prior comorbidities. Financial aid patients were defined based on their participation in the Comprehensive Social Security Assistance (CSSA) scheme, higher disability allowance, normal disability allowance, waiver and other financial aid in Hong Kong. Charlson’s standard comorbidity index and the duration and frequency of SGLT2I and DPP4I usage were calculated. To calculate the diabetes duration, the earliest date was examined by the first date of (1) diagnosis using ICD-9; (2) Hba1c≥6.5%; (3) Fasting glucose≥7.0 mmol/l or Random glucose 11.1 mmol/l; (4) using antidiabetic drugs. The estimated glomerular filtration rate (eGFR) was computed using the abbreviated modification of diet in renal disease (MDRD) formula.^16^

### Adverse outcomes

The primary outcome of this study was new-onset NPC (ICD-9: 147) and H&N cancer other than NPC (ICD-9: 140-149, 160-161, excluding 147).^17^ Mortality data were obtained from the Hong Kong Death Registry, an official government registry with the registered death records of all Hong Kong citizens linked to CDARS. Mortality was recorded according to the *International Classification of Diseases Tenth Edition* (ICD-10). The endpoint date of interest for eligible patients was the primary outcome presentation date. The endpoint for those without primary outcome was the mortality date or the endpoint of the study, which is 31^st^ December 2020.

### Statistical analysis

Descriptive statistics are used to outline the baseline clinical and biochemical characteristics of patients using SGLT2I and DPP4I. The continuous variable for baseline clinical features was expressed as the mean (95% confidence interval/standard deviation [SD]) while the total number (percentage) represented the categorical variables. The two-tailed Mann-Whitney U test was utilised to make a comparison between the continuous variables, whereas the two-tailed Chi-square test with Yates’ correction was applied for 2×2 contingency data. Propensity score matching with 1:1 ratio for SGLT2I use versus DPP4I use based on demographics, prior comorbidities, number of prior hospitalisation, duration from T2DM diagnosis initial drug exposure, non-SGLT2I/DPP4I medications, abbreviated MDRD, time-weighted HbA1c, and time-weighted fasting glucose were performed using the nearest neighbour search strategy with a calliper of 0.1. Stata software (Version 16.0) was used in facilitating the propensity score matching procedures.

In this study, we assessed the comparison of the baseline characteristics of patients before and after consuming SGLT2I and DPP4I with standardized mean difference (SMD), where SMD<0.10 was considered as indicative of well-balanced between the two groups. A Cox proportional hazards regression model was performed to identify significant risk predictors of adverse study outcomes. Adjustments were made for the demographics, past comorbidities, duration of diabetes mellitus, and number of prior hospitalizations, number of anti-diabetic drugs, non-SGLT2I/DPP4I medications, abbreviated MDRD, HbA1c, fasting glucose, lipid profile. To verify the proportionality assumption for the model mentioned above, the log-log plot was established. The cumulative incidence curves were generated for both primary and secondary outcomes and the subgroup analysis was undertaken for the affirmation of the association among patients with pivotal clinical predictors. P_interaction_ was applied to examine the differences in the associations between the subgroups.

To account for the potential competing risks in this study, cause-specific and sub-distribution hazard models were also carried out. Multiple propensity score methodologies were implemented, encompassing propensity score stratification,^18^ propensity score with inverse probability of treatment weighting (IPTW) ^19^ and propensity score with stable inverse probability weighting ^20^. We also performed a sensitivity analysis to examine the impacts of the various minimum drug duration on the outcome. The contraindication of the use of SGLT2I included patients with CKD stage 4/5 (eGFR <30 mL/min/1.73m^2), peritoneal dialysis or haemodialysis recipients and thus being excluded for the above analysis. Besides, the exclusion criteria were also included: (1) patients who have primary immunodeficiency; (2) Lag time of 6 months, 1 year and 2 years; (3) 10% propensity score at the 2 ends.

To evaluate the relationship and preference amongst the three novel second-line antidiabetic drugs, the three-arm sensitivity analysis involving GLP1a using stabilized IPTW was performed. As part of a falsification analysis to detect residual bias and confounding factors arising from unobserved variables, a negative control outcome was introduced. To be specified, lower limb fractures (ICD-9 codes: 820-829) were utilized in the falsification analysis. Any observed significant association in this context was interpreted as likely stemming from bias rather than a genuine effect. The final results were demonstrated in terms of hazard ratio (HR), 95% CI and P-value. Statistical significance was defined as P-value < 0.05. All statistical analyses were executed with RStudio software (Version: 1.1.456) and Python (Version: 3.6).

## Results

### Baseline characteristics

In this territory-wide cohort study of 76,147 patients with T2DM treated with SGLT2I/DPP4I between 1st January 2015 and 31st December 2019 in Hong Kong, patients were followed up until 31st December 2020 or their deaths (**Figure 1**). The following patient groups were excluded: (1) with prior NPC or H&N cancer (N=113); (2) without complete demographics (N=19); (3) under 18 years old (N=108). After exclusion, this study included a total of 75,884 patients with T2DM (mean age: 62.4 years old [Standard deviation: 12.8]; 55.77% males), of whom 28,778 patients used SGLT2Is, and 47,106patients used DPP4Is (**Table 1**). Before matching, the SGLT2 users had higher numbers of male patients, were younger, had lower Charlson’s standard comorbidity index, and had more cases of hypertension, use of anti-diabetic drugs, anti-hypertensive drugs and lipid-lowering drugs, and a higher eGFR compared to DPP4I users. The characteristics of patients are shown in **Table 1**.

**Table 1.** Baseline and clinical characteris2cs of pa2ents with SGLT2I v.s. DPP4I use before and aAer propensity score matching (1:1) . *for SMD≥0.1; SGLT2I: sodium glucose cotransporter-2 inhibitor; DPP4I: dipepEdyl pepEdase-4 inhibitor; MDRD: modificaEon of diet in renal disease; ACEI: angiotensin-converEng enzyme inhibitors; ARB: angiotensin II receptor blockers.

| Characteristics | Before matching |  |  |  | After matching |  |  |  |
| --- | --- | --- | --- | --- | --- | --- | --- | --- |
|  | All (N=75884)<br>Mean(SD);N or<br>Count(%) | SGLT2I users<br>(N=28778)<br>Mean(SD);N or<br>Count(%) | DPP4I users<br>(N=47106)<br>Mean(SD);N or<br>Count(%) | SMD | All (N=57556)<br>Mean(SD);N or<br>Count(%) | SGLT2I users<br>(N=28778)<br>Mean(SD);N or<br>Count(%) | DPP4I users<br>(N=28778)<br>Mean(SD);N or<br>Count(%) | SMD |
| **Demographics** |  |  |  |  |  |  |  |  |
| Male gender | 42323(55.77%) | 17127(59.51%) | 25196(53.48%) | 0.12 | 34610(60.13%) | 17127(59.51%) | 17483(60.75%) | 0.03 |
| Female gender | 33561(44.22%) | 11651(40.48%) | 21910(46.51%) | 0.12 | 22946(39.86%) | 11651(40.48%) | 11295(39.24%) | 0.03 |
| Baseline age, years | 62.4(12.8);n=75884 | 57.9(11.3);n=28778 | 65.1(12.9);n=47106 | 0.6* | 58.7(11.2);n=57556 | 57.9(11.3);n=28778 | 59.5(11.0);n=28778 | 0.14* |
| Financial aid | 2790(3.67%) | 1167(4.05%) | 1623(3.44%) | 0.03 | 2256(3.91%) | 1167(4.05%) | 1089(3.78%) | 0.01 |
| **Past comorbidities** |  |  |  |  |  |  |  |  |
| Charlson’s standard comorbidity index | 2.0(1.5);n=75884 | 1.6(1.3);n=28778 | 2.3(1.6);n=47106 | 0.53* | 1.6(1.2);n=57556 | 1.6(1.3);n=28778 | 1.7(1.2);n=28778 | 0.08 |
| Duration from earliest diabetes mellitus diagnosis date to baseline date, day | 6.5(4.9);n=75884 | 6.49(4.87);n=28778 | 6.53(4.97);n=47106 | 0.01 | 6.4(4.7);n=57556 | 6.5(4.9);n=28778 | 6.3(4.6);n=28778 | 0.05 |
| Number of hospitalizations | 1.2(0.8);n=75884 | 1.3(1.1);n=28778 | 1.2(0.6);n=47106 | 0.17 | 1.3(1.0);n=57556 | 1.3(1.1);n=28778 | 1.2(0.8);n=28778 | 0.12* |
| Hyperlipidaemia | 55285(72.85%) | 21876(76.01%) | 33409(70.92%) | 0.12 | 43800(76.09%) | 21876(76.01%) | 21924(76.18%) | <0.01 |
| Hypertension | 43161(56.87%) | 19179(66.64%) | 23982(50.91%) | 0.32* | 39318(68.31%) | 19179(66.64%) | 20139(69.98%) | 0.07 |
| Diabetic retinopathy | 5309(6.99%) | 1953(6.78%) | 3356(7.12%) | 0.01 | 3759(6.53%) | 1953(6.78%) | 1806(6.27%) | 0.02 |
| Diabetic nephropathy | 934(1.23%) | 172(0.59%) | 762(1.61%) | 0.1 | 343(0.59%) | 172(0.59%) | 171(0.59%) | <0.01 |
| Diabetic neuropathy | 1842(2.42%) | 680(2.36%) | 1162(2.46%) | 0.01 | 1343(2.33%) | 680(2.36%) | 663(2.30%) | <0.01 |
| Baseline cancer | 1940(2.55%) | 564(1.95%) | 1376(2.92%) | 0.06 | 1126(1.95%) | 564(1.95%) | 562(1.95%) | <0.01 |
| Renal diseases | 6096(8.03%) | 1536(5.33%) | 4560(9.68%) | 0.17 | 2995(5.20%) | 1536(5.33%) | 1459(5.06%) | 0.01 |
| Liver diseases | 2369(3.12%) | 1053(3.65%) | 1316(2.79%) | 0.05 | 2044(3.55%) | 1053(3.65%) | 991(3.44%) | 0.01 |
| Heart failure | 2505(3.30%) | 757(2.63%) | 1748(3.71%) | 0.06 | 1492(2.59%) | 757(2.63%) | 735(2.55%) | <0.01 |
| Acute myocardial infarction | 2158(2.84%) | 955(3.31%) | 1203(2.55%) | 0.05 | 1886(3.27%) | 955(3.31%) | 931(3.23%) | <0.01 |
| Ischemic heart disease | 7860(10.35%) | 3619(12.57%) | 4241(9.00%) | 0.12 | 6813(11.83%) | 3619(12.57%) | 3194(11.09%) | 0.05 |
| Atrial fibrillation | 1979(2.60%) | 656(2.27%) | 1323(2.80%) | 0.03 | 1300(2.25%) | 656(2.27%) | 644(2.23%) | <0.01 |
| Stroke/transient ischemic attack | 2418(3.18%) | 739(2.56%) | 1679(3.56%) | 0.06 | 1467(2.54%) | 739(2.56%) | 728(2.52%) | <0.01 |
| Autoimmune disease | 809(1.06%) | 312(1.08%) | 497(1.05%) | <0.01 | 618(1.07%) | 312(1.08%) | 306(1.06%) | <0.01 |
| HPV infection | 128(0.16%) | 45(0.15%) | 83(0.17%) | <0.01 | 90(0.15%) | 45(0.15%) | 45(0.15%) | <0.01 |
| Primary immunodeficiency | 492(0.64%) | 105(0.36%) | 387(0.82%) | 0.06 | 210(0.36%) | 105(0.36%) | 105(0.36%) | <0.01 |
| Overweight, obesity and hyperalimentation required treatment | 576(0.75%) | 421(1.46%) | 155(0.32%) | 0.12 | 827(1.43%) | 421(1.46%) | 406(1.41%) | <0.01 |
| Alcohol dependence required treatment | 169(0.22%) | 42(0.14%) | 127(0.26%) | 0.03 | 84(0.14%) | 42(0.14%) | 42(0.14%) | <0.01 |
| Tobacco dependence required treatment | 1066(1.40%) | 163(0.56%) | 903(1.91%) | 0.12 | 326(0.56%) | 163(0.56%) | 163(0.56%) | <0.01 |
| **Medications** |  |  |  |  |  |  |  |  |
| Number of anti-diabetic drug classes | 3.4(0.9);n=75884 | 3.5(1.0);n=28778 | 3.4(0.8);n=47106 | 0.2 | 3.5(0.9);n=57556 | 3.55(0.96);n=28778 | 3.55(0.91);n=28778 | <0.01 |
| Metformin | 68140(89.79%) | 26863(93.34%) | 41277(87.62%) | 0.2 | 53814(93.49%) | 26863(93.34%) | 26951(93.65%) | 0.01 |
| Sulphonylurea | 58324(76.85%) | 20695(71.91%) | 37629(79.88%) | 0.19 | 42438(73.73%) | 20695(71.91%) | 21743(75.55%) | 0.08 |
| Insulin | 38694(50.99%) | 14795(51.41%) | 23899(50.73%) | 0.01 | 29828(51.82%) | 14795(51.41%) | 15033(52.23%) | 0.02 |
| Acarbose | 2043(2.69%) | 1113(3.86%) | 930(1.97%) | 0.11 | 2135(3.70%) | 1113(3.86%) | 1022(3.55%) | 0.02 |
| Thiozolidinedone | 15218(20.05%) | 7849(27.27%) | 7369(15.64%) | 0.29* | 14751(25.62%) | 7849(27.27%) | 6902(23.98%) | 0.08 |
| Glucagon-like peptide-1 receptor agonists | 2523(3.32%) | 1967(6.83%) | 556(1.18%) | 0.29* | 3670(6.37%) | 1967(6.83%) | 1703(5.91%) | 0.04 |
| ACEI/ARB | 26842(35.37%) | 10601(36.83%) | 16241(34.47%) | 0.05 | 22617(39.29%) | 10601(36.83%) | 12016(41.75%) | 0.1 |
| Diuretics | 41874(55.18%) | 20840(72.41%) | 21034(44.65%) | 0.59* | 41039(71.30%) | 20840(72.41%) | 20199(70.18%) | 0.05 |
| Statins and fibrates | 40813(53.78%) | 18266(63.47%) | 22547(47.86%) | 0.32* | 36160(62.82%) | 18266(63.47%) | 17894(62.17%) | 0.03 |
| Systemic steroids | 434(0.57%) | 306(1.06%) | 128(0.27%) | 0.1 | 602(1.04%) | 306(1.06%) | 296(1.02%) | <0.01 |
| Immunosuppressant and chemotherapy | 2536(3.34%) | 564(1.95%) | 1972(4.18%) | 0.13 | 1132(1.96%) | 564(1.95%) | 568(1.97%) | <0.01 |
| **Calculated biomarkers** |  |  |  |  |  |  |  |  |
| Abbreviated MDRD, mL/min/1.73m<sup>2</sup> | 80.8(28.4);n=63103 | 89.8(24.1);n=24210 | 75.2(29.4);n=38893 | 0.54* | 86.6(25.8);n=48318 | 89.8(24.1);n=24210 | 86.5(27.0);n=24108 | 0.11* |
| **Lipid profiles** |  |  |  |  |  |  |  |  |
| Time weighted mean of triglyceride, mmol/L | 1.8(1.0);n=38857 | 1.9(1.0);n=17394 | 1.8(1.0);n=21463 | 0.05 | 1.9(1.0);n=32518 | 1.86(1.0);n=17394 | 1.87(1.01);n=15124 | <0.01 |
| Time weighted mean of low-density lipoprotein, mmol/L | 2.3(0.6);n=36488 | 2.3(0.56);n=16268 | 2.34(0.59);n=20220 | 0.07 | 2.3(0.6);n=30701 | 2.3(0.56);n=16268 | 2.31(0.58);n=14433 | 0.01 |
| Time weighted mean of high-density lipoprotein, mmol/L | 1.2(0.2);n=38334 | 1.16(0.19);n=17260 | 1.18(0.21);n=21074 | 0.08 | 1.2(0.2);n=32290 | 1.16(0.19);n=17260 | 1.16(0.2);n=15030 | <0.01 |
| Time weighted mean of total cholesterol, mmol/L | 4.3(0.6);n=38898 | 4.3(0.6);n=17424 | 4.4(0.6);n=21474 | 0.08 | 4.3(0.6);n=32560 | 4.3(0.6);n=17424 | 4.32(0.63);n=15136 | 0.02 |
| **Glucose tests** |  |  |  |  |  |  |  |  |
| Time weighted mean of HbA1C, % | 7.9(1.4);n=46174 | 7.9(1.4);n=19835 | 7.8(1.4);n=26339 | 0.06 | 7.9(1.4);n=38267 | 7.9(1.4);n=19835 | 7.8(1.3);n=18432 | 0.05 |
| Time weighted mean of fasting glucose, mmol/L | 7.8(3.0);n=42839 | 7.9(2.9);n=18576 | 7.8(3.0);n=24263 | 0.03 | 7.9(3.1);n=35309 | 7.9(2.9);n=18576 | 7.8(3.2);n=16733 | 0.02 |

After the propensity score matching, baseline characteristics and the time-weighted lipid and glucose profiles of the 2 groups were well-balanced, apart from baseline age (SMD=0.14), number of hospitalizations (SMD=0.12), thiazolidinedione (SMD=0.13), and abbreviated MDRD (SMD=0.11) **(Table 1)**. The DPP4I and SGLT2I cohorts were comparable after matching with nearest neighbour search strategy with calliper of 0.1, and the proportional hazard assumption was confirmed **(Supplementary** Figure 1). In the matched cohort, 106 patients developed NPC, 50 patients developed H&N cancer, 899 patients passed away due to cancer during the study period **(Figure 1)**.

### The association between SGLT2I and the primary outcomes

In the matched cohort, 31 SGLT2I users and 75 DPP4I users developed NPC. After a mean follow-up of 314,709.1 person-year, the incidence of NPC was lower amongst SGLT2I users (Incidence rate [IR] per 100,000 person-year: 19.43; 95% CI: 13.20-27.58) compared to DPP4I users (IR per 100,000 person-year: 48.33; 95% CI: 38.02-60.58) **(Table 2)**. Compared to DPP4I users, SGLT2I users had a 59% lower risk of NPC after adjustment (Hazard ratio [HR]: 0.41; 95% Confidence Interval [CI]: 0.21-0.81) for demographics, past comorbidities, duration of diabetes mellitus, and number of prior hospitalizations, number of anti-diabetic drugs, non-SGLT2I/DPP4I medications, abbreviated MDRD, HbA1c, fasting glucose, lipid profile. **(Table 2)**. This was substantiated by the cumulative incidence curves stratified by SGLT2I versus DPP4I **(Figure 2)**.

**Figure 2.**
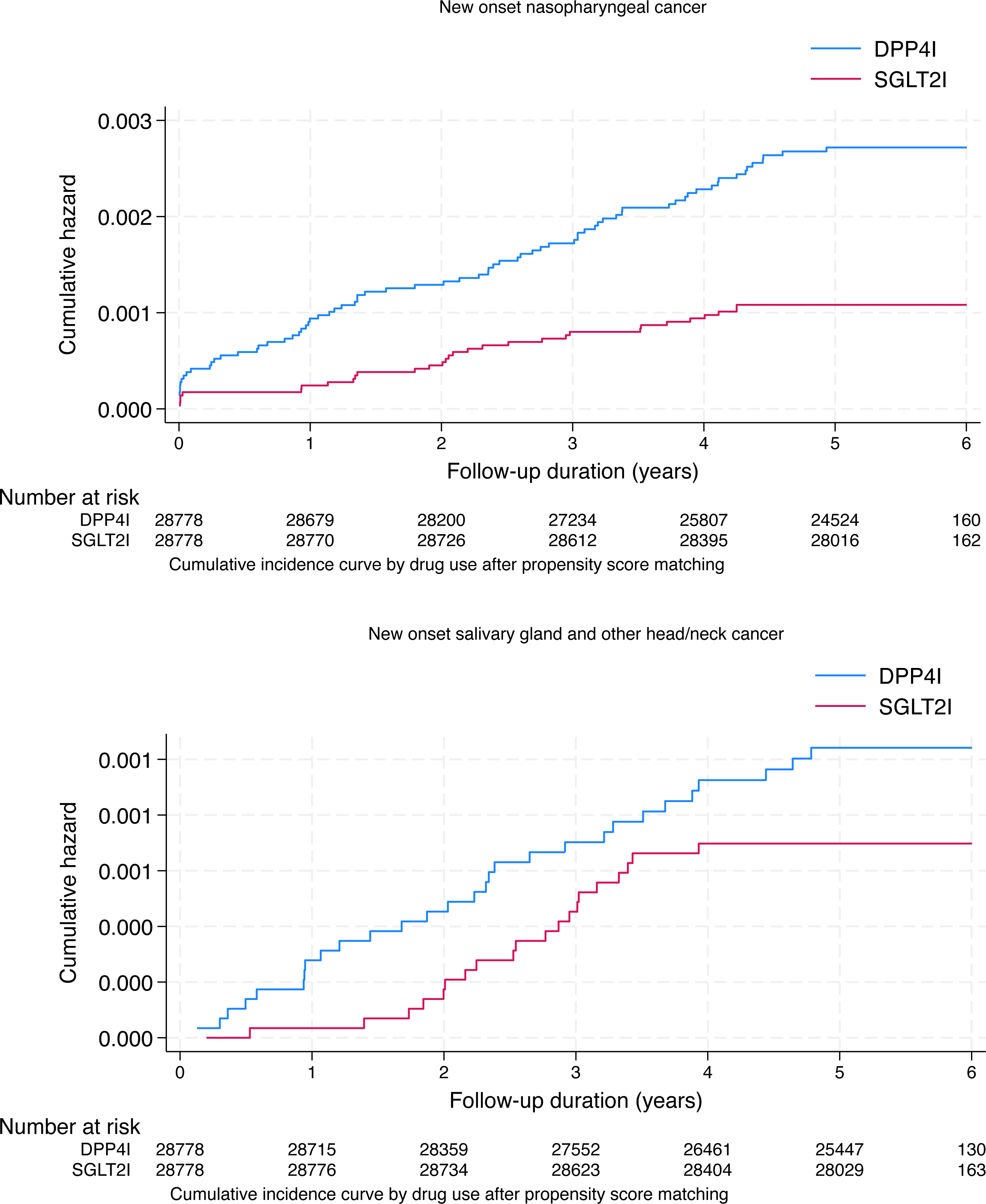
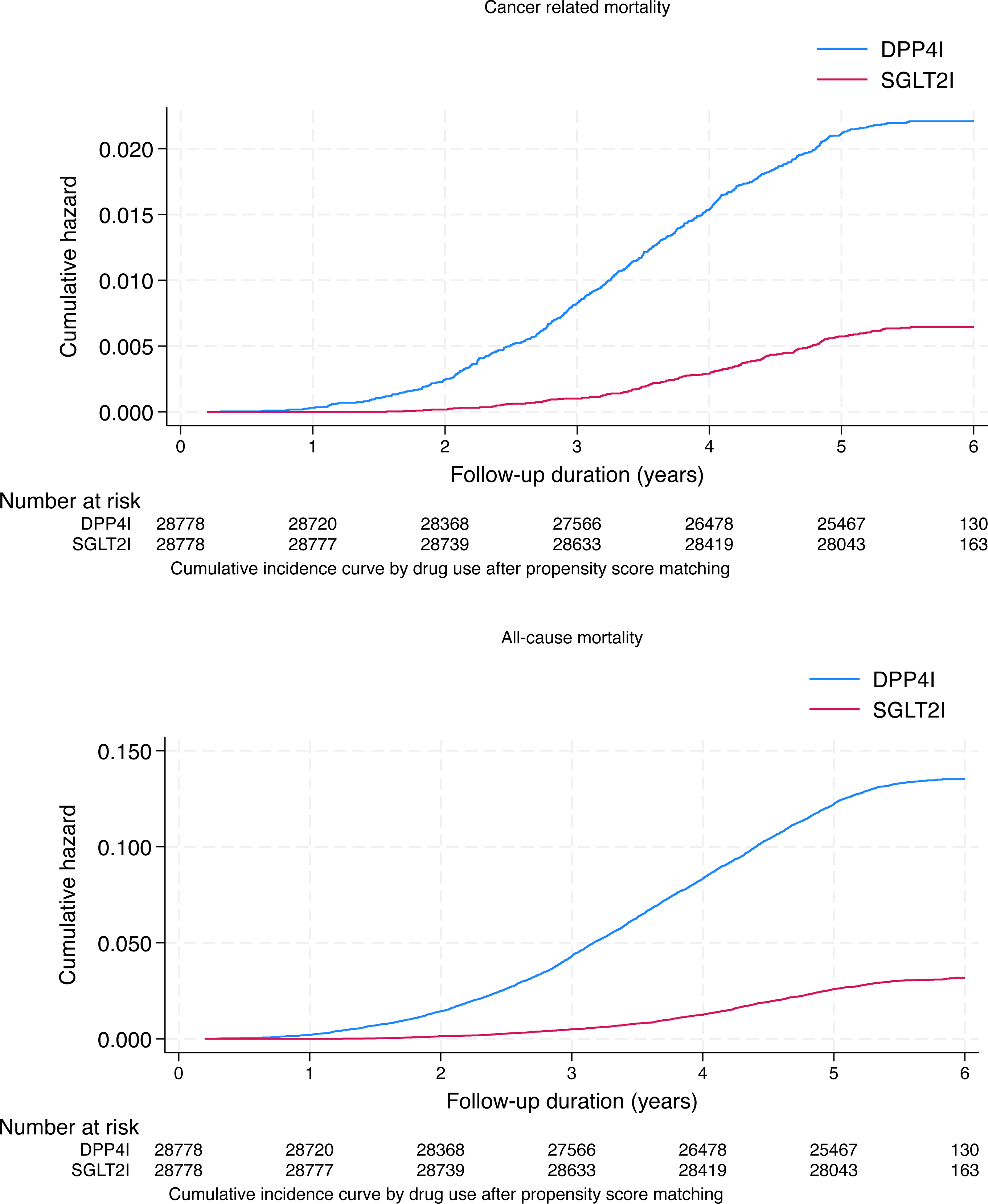
Cumulative incidence curves for new onset NPC, other H&N cancer, cancer-related mortality, and all-cause mortality stratified by drug exposure of SGLT2I and DPP4I after propensity score matching (1:1) SGLT2I: Sodium-glucose cotransporter-2 inhibitors; DPP4I: Dipeptidyl peptidase-4 inhibitors. NPC: nasopharyngeal carcinoma; H&N: head and neck.

**Table 2.**
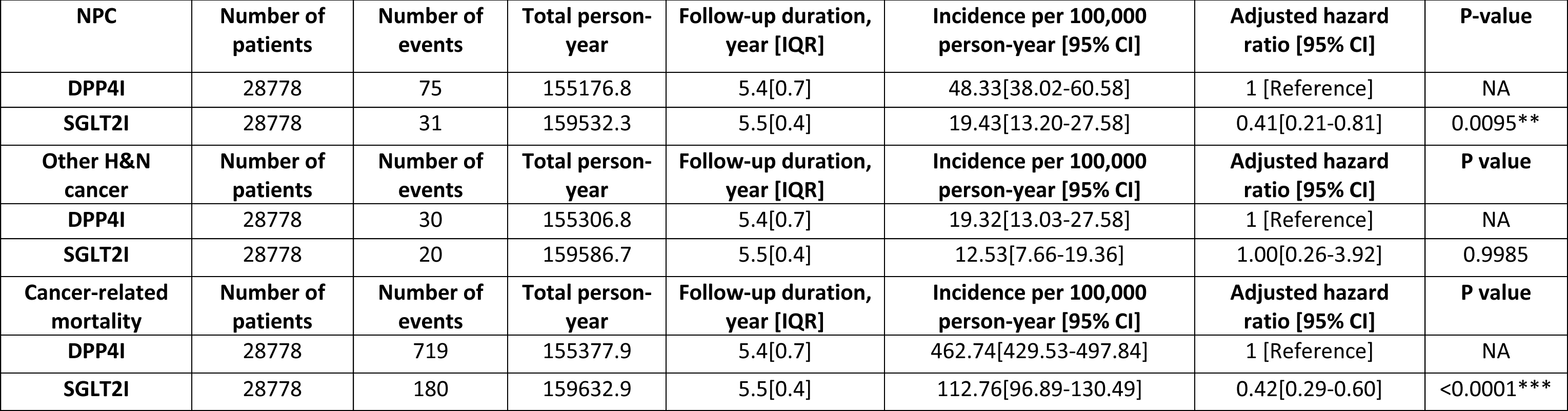
Incidence and mul2variate Cox regression models of new onset NPC, other H&N cancer and cancer-related mortality in the cohort after 1:1 propensity score matching. . *For p≤ 0.05, ** for p ≤ 0.01, *** for p ≤ 0.001; CI: confidence interval; SGLT2I: sodium glucose cotransporter-2 inhibitor; DPP4I: dipeptidyl peptidase-4 inhibitor; IQR: Interquartile range. Adjusted for significant demographics, past comorbidities, duration of diabetes mellitus, and number of prior hospitalizations, number of anti-diabetic drugs, non-SGLT2I/DPP4I medications, abbreviated MDRD, HbA1c, fasting glucose, lipid profile.

| NPC | Number of patients | Number of events | Total person-year | Follow-up duration, year [IQR] | Incidence per 100,000 person-year [95% CI] | Adjusted hazard ratio [95% CI] | P-value |
| --- | --- | --- | --- | --- | --- | --- | --- |
| DPP4I | 28778 | 75 | 155176.8 | 5.4[0.7] | 48.33[38.02-60.58] | 1 [Reference] | NA |
| SGLT2I | 28778 | 31 | 159532.3 | 5.5[0.4] | 19.43[13.20-27.58] | 0.41[0.21-0.81] | 0.0095** |
| Other H&N cancer | Number of patients | Number of events | Total person-year | Follow-up duration, year [IQR] | Incidence per 100,000 person-year [95% CI] | Adjusted hazard ratio [95% CI] | P value |
| DPP4I | 28778 | 30 | 155306.8 | 5.4[0.7] | 19.32[13.03-27.58] | 1 [Reference] | NA |
| SGLT2I | 28778 | 20 | 159586.7 | 5.5[0.4] | 12.53[7.66-19.36] | 1.00[0.26-3.92] | 0.9985 |
| Cancer-related mortality | Number of patients | Number of events | Total person-year | Follow-up duration, year [IQR] | Incidence per 100,000 person-year [95% CI] | Adjusted hazard ratio [95% CI] | P value |
| DPP4I | 28778 | 719 | 155377.9 | 5.4[0.7] | 462.74[429.53-497.84] | 1 [Reference] | NA |
| SGLT2I | 28778 | 180 | 159632.9 | 5.5[0.4] | 112.76[96.89-130.49] | 0.42[0.29-0.60] | <0.0001*** |

Meanwhile, 20 SGLT2I users and 30 DPP4I users developed H&N cancer. After a mean follow-up of 314,893.5 person-year, the incidence of NPC was lower amongst SGLT2I users (IR per 100,000 person-year: 12.53; 95% CI: 7.66-19.36) compared to DPP4I users (IR per 100,000 person-year: 19.32; 95% CI: 13.03-27.58) **(Table 2)**. SGLT2I users was not associated with H&N cancer (HR: 1.00; 95% CI: 0.26-3.92) compared to users of DPP4I after adjustment **(Table 2)**.

### The association between SGLT2I and cancer-related mortality

180 SGLT2I users and 719 DPP4I users died. After a follow-up of 315,010.8 person-year, the incidence of cancer-related mortality was lower amongst SGLT2I users (IR: 112.76; 95% CI: 96.89-130.49) compared to DPP4I users (IR: 462.74; 95% CI: 429.53-497.84) **(Table 2)**. SGLT2I users had a lower risk of cancer-related mortality after adjustment (HR: 0.42; 95% CI: 0.29-0.60) compared to users of DPP4I. This was substantiated by the cumulative incidence curves stratified by SGLT2I versus DPP4I **(Figure 2)**.

### Subgroup analysis

The results of the subgroup analysis for effects of SGLT2I and DPP4I on the NPC are shown in **Figure 3**. The result demonstrated that SGLT2I was associated with lower risks of NPC regardless of sex, age, socioeconomic status, renal disease, and obesity. Besides, the effects of SGLT2I on NPC were not affected by presence of baseline cancer, autoimmune disease, systemic steroid use, or immunosuppressant/chemotherapy use. The marginal effects analysis demonstrated that SGLT2I use was associated with lower risks of NPC regardless the duration of diabetes (**Supplementary** Figure 2). However, SGLT2I was associated with lower risks of NPC compared to DPP4I only amongst patient with poorer renal function and higher time-weighted mean of HbA1c. Meanwhile, SGLT2I was associated with lower risks of H&N cancer regardless of the time-weighted mean of HbA1c, but only amongst patients with short duration of diabetes, poorer renal function.

**Figure 3.**
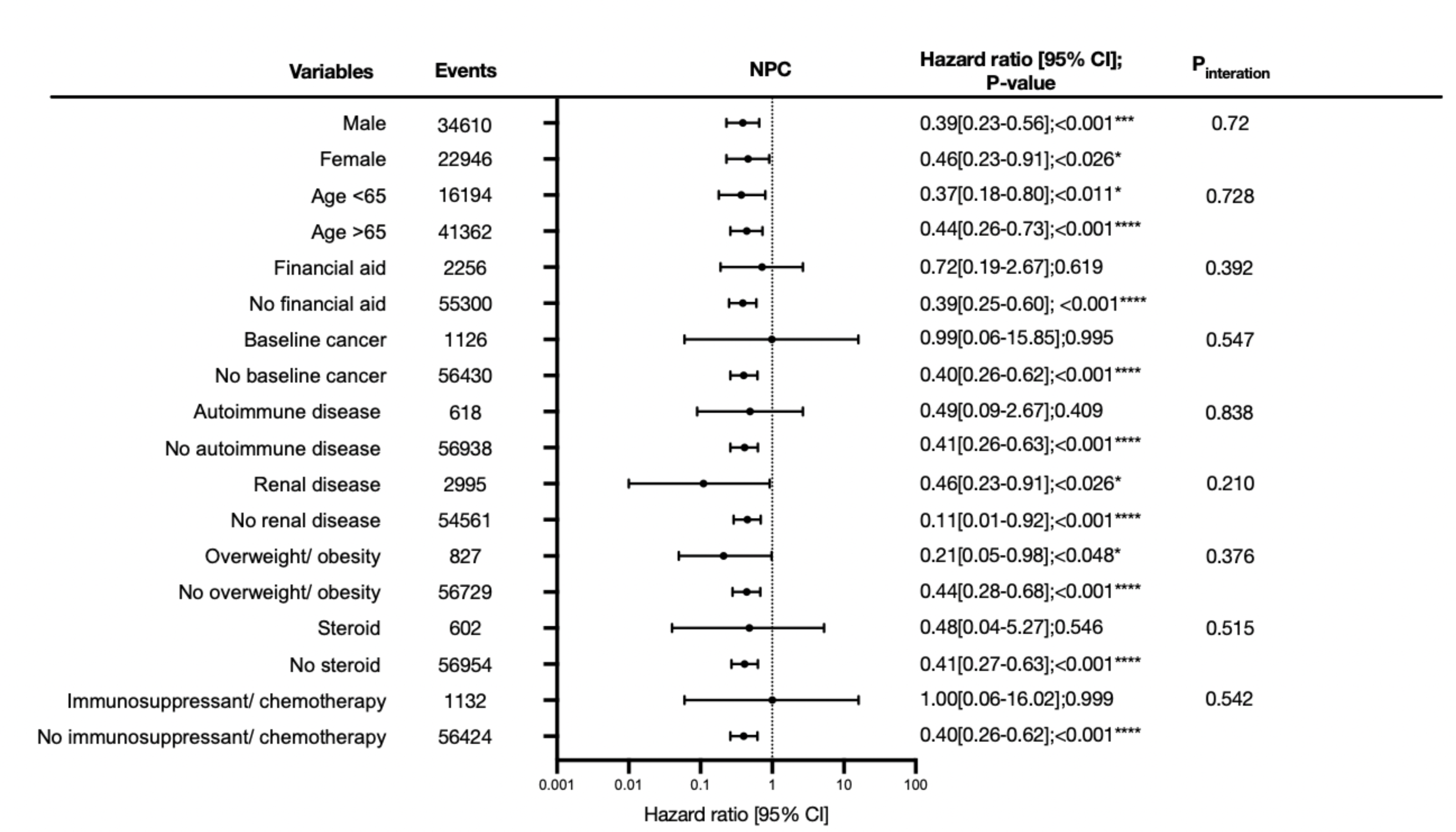
**Subgroup analyses for SGLT2I v.s. DPP4I exposure predict new onset NPC in the matched cohort**. For p≤ 0.05, ** for p ≤ 0.01, *** for p ≤ 0.001; CI: confidence interval; SGLT2I: Sodium-glucose cotransporter-2 inhibitors; DPP4I: Dipeptidyl peptidase-4 inhibitors; HR: Hazard ratio; NPC: nasopharyngeal carcinoma.

### Sensitivity analyses

Sensitivity analyses were performed to confirm the predictability of the models. The results of the cause-specific hazard models, sub-distribution hazard models and different propensity score approaches demonstrated that different models did not change association for both the primary and the secondary outcomes (**Supplementary Table 2)**. A three-arm analysis with the inclusion of GLP1a including patients only on (SGLT2I, DPP4I, and GLP1a) using stabilized IPTW was conducted **(Supplementary Table 3)**. While SGLT2I was also associated with higher risks of NPC compared to GLP1a, the results remained insignificant due to the insufficient power in the GLP1a cohort (HR: 1.24; 95% CI: 0.79-1.66). Besides, SGLT2I also did not associated with a lower risk of cancer-related mortality compared to GLP1a (HR: 1.09; 95% CI: 0.56-1.95). The sensitivity analysis for 6 months, 1-year and 2-year lag time demonstrated the same pattern. Besides, after excluding patients with extreme 10% propensity score at the 2 end, patients with CKD stage 4/5 (eGFR <30), peritoneal dialysis or haemodialysis, and patients with primary immunodeficiency likewise, the results demonstrated that SGLT2I remained associated with lower risks of NPC compared to DPP4I **(Supplementary Table 4)**.

### Falsification analysis

Lower limb fracture was used as negative control outcome in the falsification analysis for the comparison between SGLT2I and DPP4I since most of the cases should be traumatic cause **(Supplementary Table 9)**. The results demonstrated that SGLT2I was not associated with lower limb fracture after adjustment (HR: 1.29; 95% CI: 00.95-1.79) compared to DPP4I.

## Discussion

In this territory-wide cohort study, the associations between SGLT2I, DPP4I and new-onset NPC and H&N were examined using real-world data. Our results firstly demonstrate that SGLT2I use was independently associated with a lowered risk of new-onset NPC than DPP4I usage after adjustments. Secondly, SGLT2I was not associated with lower risks of H&N cancer compared to DPP4I. The results remained consistent across different competing risk and the sensitivity analyses, and the falsification analysis did not falsify the results. To the best of our knowledge, this was the first cohort study to examine the association between SGLT2I and NPC.

### Comparison with previous studies

The incidence of NPC in our study for SGLT2I and for DPP4I parallels the values of existing study, with a reported incidence of 15-50 per 100,000 population.^1^ NPC is endemic in Southern China, as well as other parts of South-East Asia.^21^ In Hong Kong, a previous study described an incidence of 10.1 per 100,000 in male and 3.3 per 100,000 in female in 2020.^22^ Previously, SGLT2I was proposed as an potential anti-tumour agents since sodium-glucose cotransporter 2 was over-expressed in multiple tumour models, including head and neck cancer.^23, 24^ In this study, we found that SGLT2I was associated with lower risk of NPC compared to DPP4I.

Previously, metformin was reported to be associated with a lower risk of NPC compared to metformin non-user.^7, 25, 26^ However, the effects of the anti-diabetic drugs on NPC might be independent of their ability of glycaemic control. Previously, a meta-analysis identified that T2DM patients might have decreased risk of NPC amongst the Asian population. This could be explained by the pathological phenotypes of undifferentiated NPC. It was found that the prevalence of T2DM in differentiated NPC was higher compared to undifferentiated cases, such that T2DM may only effect on the former pathological subtype.^27^ T2DM was also reported not to be associated with mortality amongst NPC patients.^28^ Meanwhile, in a prospective cohort study in Taiwan, diabetic patients who received metformin had a lower risk of developing H&N cancer compared to the matched individuals without metformin. Specifically, the incidence of oropharyngeal cancer and NPC was significantly lower for patients treated with metformin than their counterparts.^29^ Another study also supported a reduced risk of H&N cancer for metformin, particularly with a history of smoking and heavy drinking.^29^ Apart from anti-diabetic medications, a Taiwan case-control study found that the risk of head and neck cancer is reduced by statin.^30^ Several studies also demonstrated that patients received statin were associated with increased survival of head and neck cancer.^31, 32^ Hence, metabolic drugs might potentially be preventive for head and neck cancer.

### Potential underlying mechanisms

SGLT2 inhibitors have recently reported to exhibit possible anti-cancer effects in several malignancies. It was proposed that SGLT2I act to inhibit the active reverse transport of glucose by SGLT-2 located in the proximal renal tubule, which consequently affect the cellular respiration and growth of the tumour cells.^33, 34^ Our results suggested that the effects of SGLT2I mainly targeted specifically on NPC but not H&N cancer in general. Besides, SGLT2I may act on the LMP1 induced glucose transporter 1 which reduce glucose uptake by the NPC cells.^6, 11^ Meanwhile, DPP4I were hypothesized to diminish the cancer risk by eliminating the degradation of incretins, glucagon-like peptide-1 (GLP-1) and glucose-dependent insulinotropic peptide (GIP) in the cell.^35^ Although there are multiple studies regarding both SGLT2 inhibitors and DPP4 inhibitors act on cancer, the mechanism of both medications towards head and neck cancer, especially NPC, remains unclear.

### Clinical implications

The secondary protective benefits of SGLT2I and DPP4I on cardiovascular benefits and cancer have received worldwide attention in recent years. While there is substantial evidence supporting the effects of the anti-diabetic medications on cardiovascular diseases ^36^, few evidence surrounding the latter. By exploring the association of SGLT2I and DPP4I in HCCNPC this study added to the growing body of evidence supporting the use of antidiabetic agents in preventing cancer. However, further molecular studies and clinical studies are needed to confirm the causation relationship between SGLT2I and NPC.

### Limitations

Several limitations should be acknowledged in this study. First of all, the observational nature of this study gives inherent issues such as under-coding, coding errors, and missing data possibly resulting in information bias. Crucial data on predictive variables, for instance, nitrosamine consumption, diet, and family history of NPC were unavailable from CDARS. In addition, the inclusion of patients in CDARS was only limited to those who smoked, consumed alcohol, and were obese to a degree that required medical attention and treatment. To mitigate this limitation, this study incorporated extensive comorbidities related to NPC to indirectly infer the possible risk variables. Furthermore, a falsification analysis was also conducted to minimise the risk of residual confounding. Moreover, the retrospective design allows only for the inference of associations but not the demonstration of causation relationships. Lastly, assessing medication adherence relied solely on prescription refills, lacking direct measurement of drug exposure. This approach may lead to time lag biases and immortal time issues. Hence, it is essential to conduct prospective randomised controlled trials aimed at evaluating the causal links of anti-diabetic medications.

## Conclusion

In this population-based cohort study, SGLT2I users was associated with lower risks of NPC but not H&N cancer compared to DPP4I users after matching and adjustments compared to DPP4I users in Asia. Future studies are needed about the SGLT2I and its action on NPC.

## Ethical approval statement

This study was approved by the Institutional Review Board of the University of Hong Kong/Hospital Authority Hong Kong West Cluster (HKU/HA HKWC IRB) (UW-20-250) and complied with the Declaration of Helsinki.

## Availability of data and materials

The data that support the findings of this study were provided by the Hong Kong Hospital Authority, but restrictions apply to the availability of these data, which were used under license for the current study, and so are not publicly available. Data are however available from the authors upon reasonable request and with permission of Hong Kong Hospital Authority.

## Acknowledgements

Oscar Hou In Chou and Kar Kei Mak would like to thank Bai Xian Asia Institute (BXAI) for her generosity in providing Asian Future Leaders Scholarship Program scholarship.

## Guarantor Statement

All authors approved the final version of the manuscript. GT is the guarantor of this work and, as such, had full access to all the data in the study and takes responsibility for the integrity of the data and the accuracy of the data analysis.

## Author contributions

LF L, OHI C, JD Z: conception of study, preparation of figures, study design, data contribution, statistical analysis, data interpretation, manuscript drafting, and critical revision of the manuscript.

OHI C, KK M, YF Y, CT C: literature search, data interpretation, literature search, data collection, manuscript drafting.

GL L, C C, WT W, T L, BMY C: critical revision of the manuscript

G T, JD Z: conception of study and literature search, study design, data collection, and critical revision of manuscript, study supervision.

## References

1. Mahdavifar N, Ghoncheh M, Mohammadian-Hafshejani A, Khosravi B, Salehiniya H. Epidemiology and Inequality in the Incidence and Mortality of Nasopharynx Cancer in Asia. Osong Public Health and Research Perspectives. 2016;7(6): 360–372. 10.1016/j.phrp.2016.11.002.

2. Getz KR, Bellile E, Zarins KR, et al. Statin use and head and neck squamous cell carcinoma outcomes. Int J Cancer. 2021;148(10): 2440–2448. 10.1002/ijc.33441.

3. Chang ET, Adami HO. The enigmatic epidemiology of nasopharyngeal carcinoma. Cancer Epidemiol Biomarkers Prev. 2006;15(10): 1765–1777. 10.1158/1055-9965.Epi-06-0353.

4. Yu MC, Yuan JM. Epidemiology of nasopharyngeal carcinoma. Semin Cancer Biol. 2002;12(6): 421–429. 10.1016/s1044579x02000858.

5. Tsao SW, Yip YL, Tsang CM, et al. Etiological factors of nasopharyngeal carcinoma. Oral Oncol. 2014;50(5): 330–338. 10.1016/j.oraloncology.2014.02.006.

6. Liu Q, Bode AM, Chen X, Luo X. Metabolic reprogramming in nasopharyngeal carcinoma: Mechanisms and therapeutic opportunities. Biochimica et Biophysica Acta (BBA) – Reviews on Cancer. 2023;1878(6): 189023. 10.1016/j.bbcan.2023.189023.

7. Tseng C-H. Metformin and risk of developing nasopharyngeal cancer in patients with type 2 diabetes mellitus. Metabolism. 2018;85: 223–226. 10.1016/j.metabol.2018.04.009.

8. Benedetti R, Benincasa G, Glass K, et al. Effects of novel SGLT2 inhibitors on cancer incidence in hyperglycemic patients: a meta-analysis of randomized clinical trials. Pharmacological Research. 2022;175: 106039. 10.1016/j.phrs.2021.106039.

9. Oscar Hou In C, Jing N, Raymond Ngai Chiu C, et al. Lower risks of sodium glucose cotransporter 2 (SGLT2) inhibitors compared to dipeptidyl peptidase-4 (DPP4) inhibitors for new-onset non-alcoholic fatty liver disease and hepatocellular carcinoma in type 2 diabetes mellitus: A population-based study. medRxiv. 2022: 2022.2008.2016.22278847. 10.1101/2022.08.16.22278847.

10. Chung CT, Lakhani I, Chou OHI, et al. Sodium-glucose cotransporter 2 inhibitors versus dipeptidyl peptidase 4 inhibitors on new-onset overall cancer in Type 2 diabetes mellitus: A population-based study. Cancer Med. 2023;12(11): 12299–12315. 10.1002/cam4.5927.

11. Zhang J, Jia L, Lin W, et al. Epstein-Barr Virus-Encoded Latent Membrane Protein 1 Upregulates Glucose Transporter 1 Transcription via the mTORC1/NF-κB Signaling Pathways. J Virol. 2017;91(6). 10.1128/jvi.02168-16.

12. Rudrapatna VA, Glicksberg BS, Butte AJ. Utility of routinely collected electronic health records data to support effectiveness evaluations in inflammatory bowel disease: a pilot study of tofacitinib. BMJ Health Care Inform. 2021;28(1). 10.1136/bmjhci-2021-100337.

13. Gao X, Zhang N, Lu L, et al. New-onset syncope in diabetic patients treated with sodium-glucose cotransporter-2 inhibitors versus dipeptidyl peptidase-4 inhibitors: a Chinese population-based cohort study. European Heart Journal – Cardiovascular Pharmacotherapy. 2023: pvad086. 10.1093/ehjcvp/pvad086.

14. Chou OHI, Zhou J, V Mui J, et al. Lower risks of new-onset acute pancreatitis and pancreatic cancer in sodium glucose cotransporter 2 (SGLT2) inhibitors compared to dipeptidyl peptidase-4 (DPP4) inhibitors: A propensity score-matched study with competing risk analysis. Diabetes Epidemiology and Management. 2023;9: 100115. 10.1016/j.deman.2022.100115.

15. Lee TTL, Chan SCL, Chou OHI, et al. Initiation of warfarin is associated with decreased mortality in patients with infective endocarditis: A population-based cohort study. Thromb Res. 2023;233: 1–9. 10.1016/j.thromres.2023.11.009.

16. Soliman AR, Fathy A, Khashab S, Shaheen N. Comparison of abbreviated modification of diet in renal disease formula (aMDRD) and the Cockroft-Gault adjusted for body surface (aCG) equations in stable renal transplant patients and living kidney donors. Ren Fail. 2013;35(1): 94–97. 10.3109/0886022x.2012.731970.

17. Chen MC, Feng IJ, Lu CH, et al. The incidence and risk of second primary cancers in patients with nasopharyngeal carcinoma: a population-based study in Taiwan over a 25-year period (1979-2003). Ann Oncol. 2008;19(6): 1180–1186. 10.1093/annonc/mdn003.

18. Austin PC. An Introduction to Propensity Score Methods for Reducing the Effects of Confounding in Observational Studies. Multivariate Behav Res. 2011;46(3): 399–424. 10.1080/00273171.2011.568786.

19. Austin PC, Stuart EA. Moving towards best practice when using inverse probability of treatment weighting (IPTW) using the propensity score to estimate causal treatment effects in observational studies. Stat Med. 2015;34(28): 3661–3679. 10.1002/sim.6607.

20. Avagyan V, Vansteelandt S. Stable inverse probability weighting estimation for longitudinal studies. Scandinavian Journal of Statistics. 2021;48(3): 1046–1067. 10.1111/sjos.12542.

21. Chen YP, Chan ATC, Le QT, Blanchard P, Sun Y, Ma J. Nasopharyngeal carcinoma. Lancet. 2019;394(10192): 64-80. 10.1016/s0140-6736(19)30956-0.

22. Wang X, Sun H, Li L, Gan Z, Wu X, Du J. Changing patterns of nasopharyngeal carcinoma incidence in Hong Kong: a 30-year analysis and future projections. BMC Cancer. 2023;23(1): 761. 10.1186/s12885-023-11296-1.

23. Basak D, Gamez D, Deb S. SGLT2 Inhibitors as Potential Anticancer Agents. Biomedicines. 2023;11(7). 10.3390/biomedicines11071867.

24. Wang Y, Mang X, Li D, et al. Cold atmospheric plasma sensitizes head and neck cancer to chemotherapy and immune checkpoint blockade therapy. Redox Biology. 2024;69: 102991. 10.1016/j.redox.2023.102991.

25. Li H, Chen X, Yu Y, et al. Metformin inhibits the growth of nasopharyngeal carcinoma cells and sensitizes the cells to radiation via inhibition of the DNA damage repair pathway. Oncol Rep. 2014;32(6): 2596–2604. 10.3892/or.2014.3485.

26. Yen YC, Lin C, Lin SW, Lin YS, Weng SF. Effect of metformin on the incidence of head and neck cancer in diabetics. Head Neck. 2015;37(9): 1268–1273. 10.1002/hed.23743.

27. Zucchetto A, Taborelli M, Bosetti C, et al. Metabolic disorders and the risk of nasopharyngeal carcinoma: a case-control study in Italy. Eur J Cancer Prev. 2018;27(2): 180–183. 10.1097/cej.0000000000000286.

28. Peng H, Chen L, Zhang Y, et al. Prognostic value of Diabetes in Patients with Nasopharyngeal Carcinoma Treated with Intensity-Modulated Radiation Therapy. Scientific Reports. 2016;6(1): 22200. 10.1038/srep22200.

29. Figueiredo RA, Weiderpass E, Tajara EH, et al. Diabetes mellitus, metformin and head and neck cancer. Oral Oncol. 2016;61: 47–54. 10.1016/j.oraloncology.2016.08.006.

30. Kao LT, Hung SH, Kao PF, Liu JC, Lin HC. Inverse association between statin use and head and neck cancer: Population-based case-control study in Han population. Head Neck. 2019;41(5): 1193–1198. 10.1002/hed.25501.

31. Gupta A, Stokes W, Eguchi M, et al. Statin use associated with improved overall and cancer specific survival in patients with head and neck cancer. Oral Oncol. 2019;90: 54–66. 10.1016/j.oraloncology.2019.01.019.

32. Lebo NL, Griffiths R, Hall S, Dimitroulakos J, Johnson-Obaseki S. Effect of statin use on oncologic outcomes in head and neck squamous cell carcinoma. Head Neck. 2018;40(8): 1697–1706. 10.1002/hed.25152.

33. Dutka M, Bobiński R, Francuz T, et al. SGLT-2 Inhibitors in Cancer Treatment-Mechanisms of Action and Emerging New Perspectives. Cancers (Basel). 2022;14(23). 10.3390/cancers14235811.

34. Nasiri AR, Rodrigues MR, Li Z, Leitner BP, Perry RJ. SGLT2 inhibition slows tumor growth in mice by reversing hyperinsulinemia. Cancer & Metabolism. 2019;7(1): 10. 10.1186/s40170-019-0203-1.

35. Zhao M, Chen J, Yuan Y, et al. Dipeptidyl peptidase-4 inhibitors and cancer risk in patients with type 2 diabetes: a meta-analysis of randomized clinical trials. Scientific Reports. 2017;7(1): 8273. 10.1038/s41598-017-07921-2.

36. McGuire DK, Shih WJ, Cosentino F, et al. Association of SGLT2 Inhibitors With Cardiovascular and Kidney Outcomes in Patients With Type 2 Diabetes: A Meta-analysis. JAMA Cardiol. 2021;6(2): 148–158. 10.1001/jamacardio.2020.4511.

