## Supplementary Appendix for "Nasopharyngeal Carcinoma and Head and Neck Cancer in Patients with Type-2 Diabetes Mellitus receiving SGLT2I, DPP4I or GLP1a: A population-based cohort study"

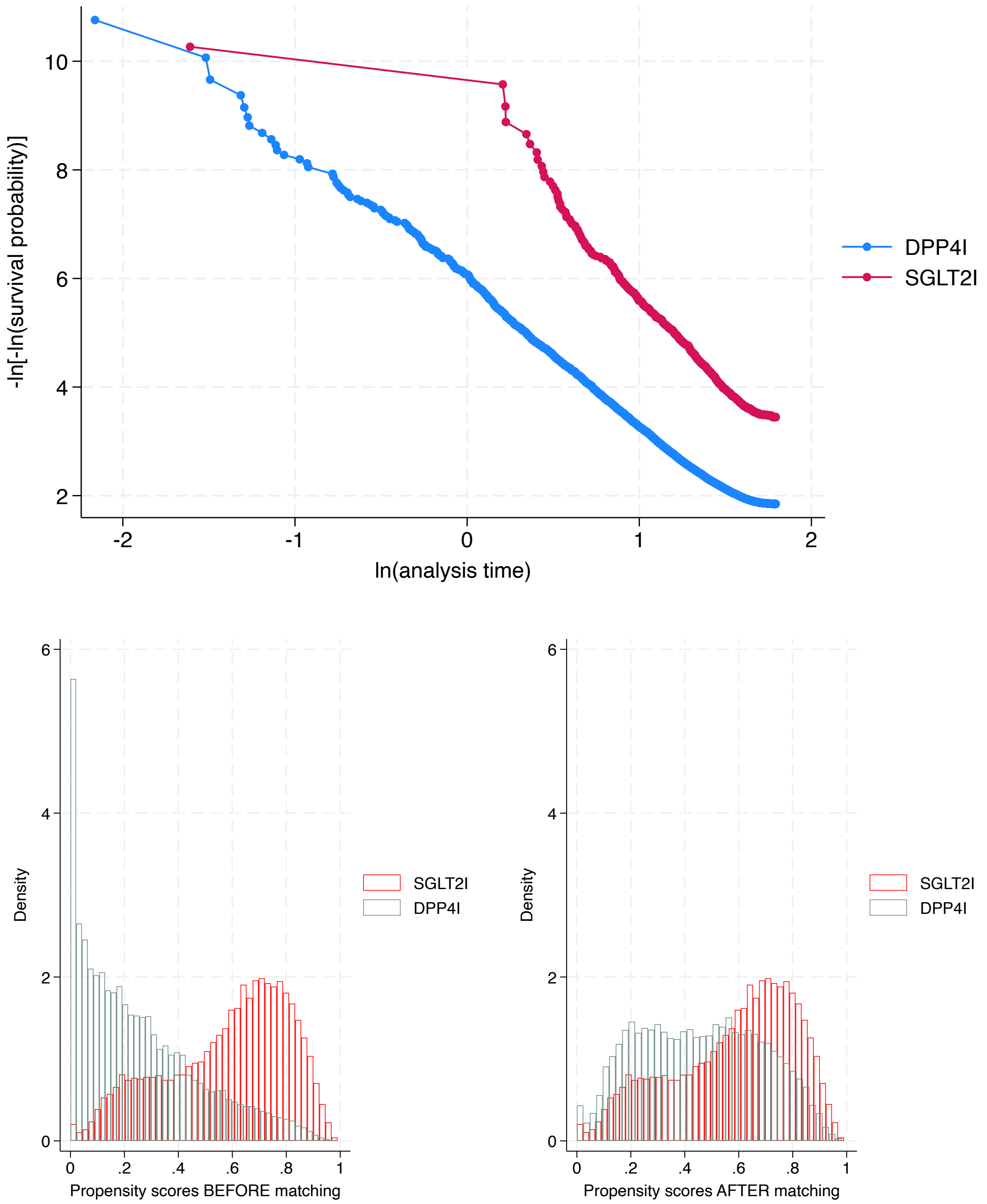

**Supplementary Figure 1. Propensity score matching comparisons and proportional hazard assumption checking with parallel lines for SGLT2I versus DPP4I before and after 1:1 matching with nearest neighbours search strategy with calliper of 0.1.**

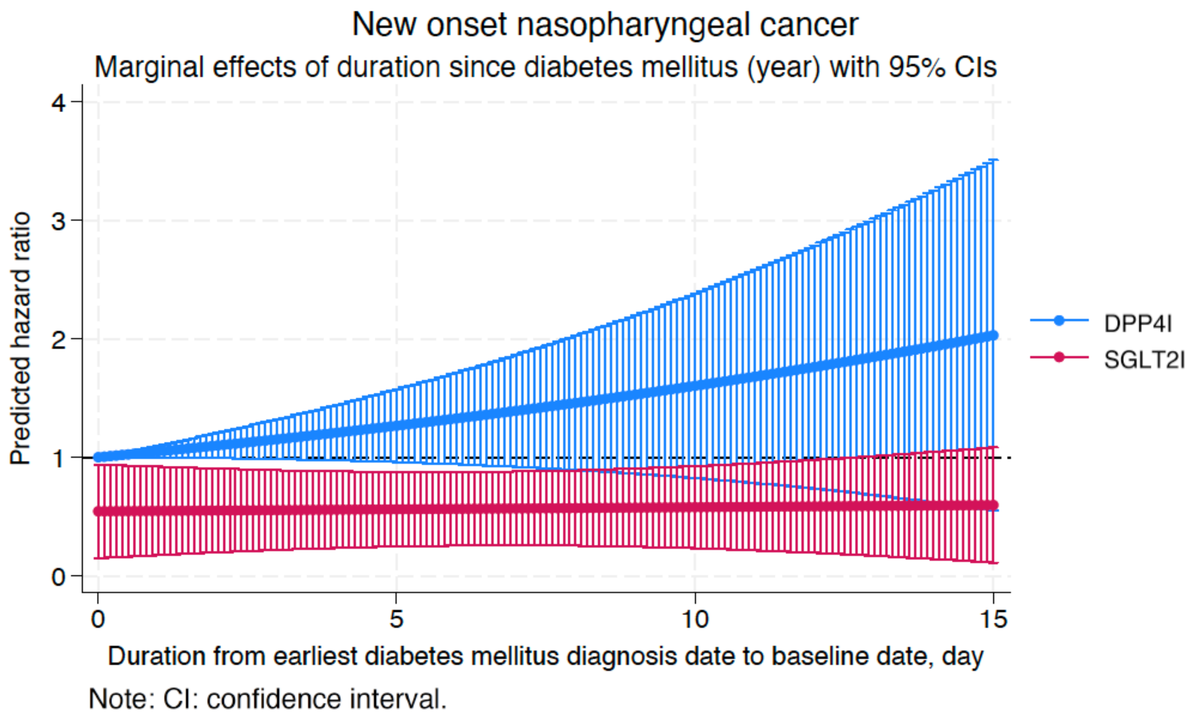
New onset nasopharyngeal cancer
Marginal effects of duration since diabetes mellitus (year) with 95% CIs
Predicted hazard ratio
0
1
2
3
4
0
5
10
15
Duration from earliest diabetes mellitus diagnosis date to baseline date, day
DPP4I
SGLT2I
Note: CI: confidence interval.

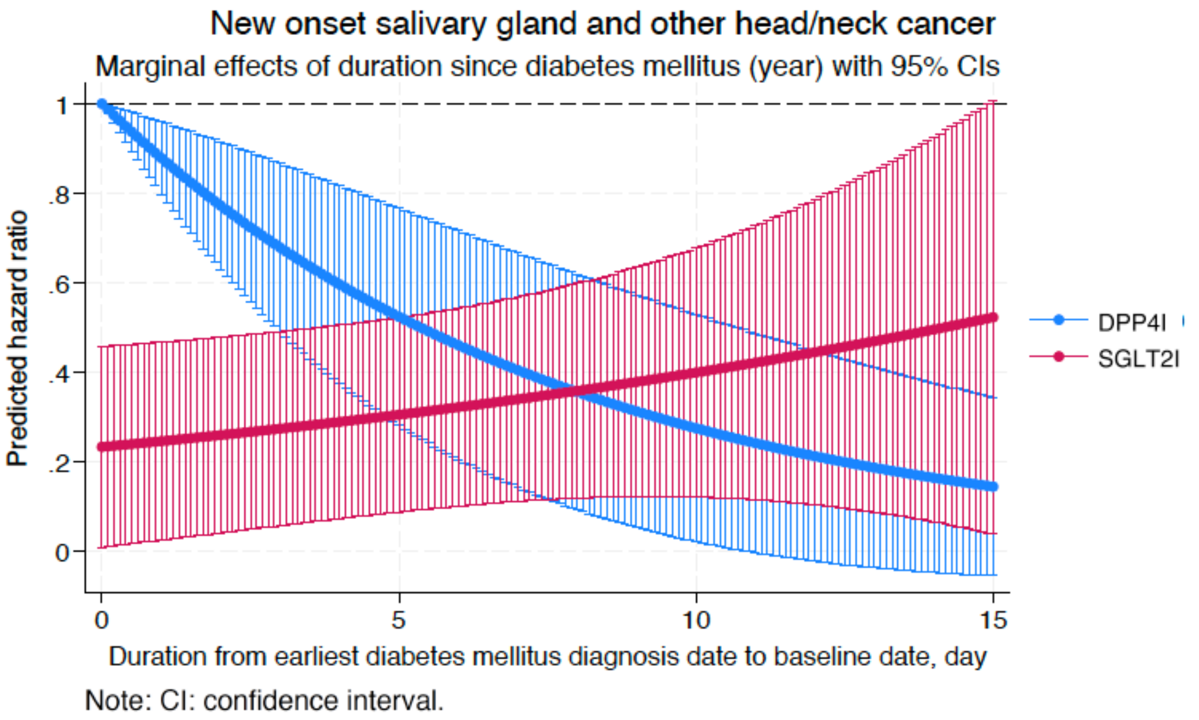
New onset salivary gland and other head/neck cancer
Marginal effects of duration since diabetes mellitus (year) with 95% CIs
Predicted hazard ratio
0
.2
.4
.6
.8
1
0
5
10
15
Duration from earliest diabetes mellitus diagnosis date to baseline date, day
DPP4I
SGLT2I
Note: CI: confidence interval.

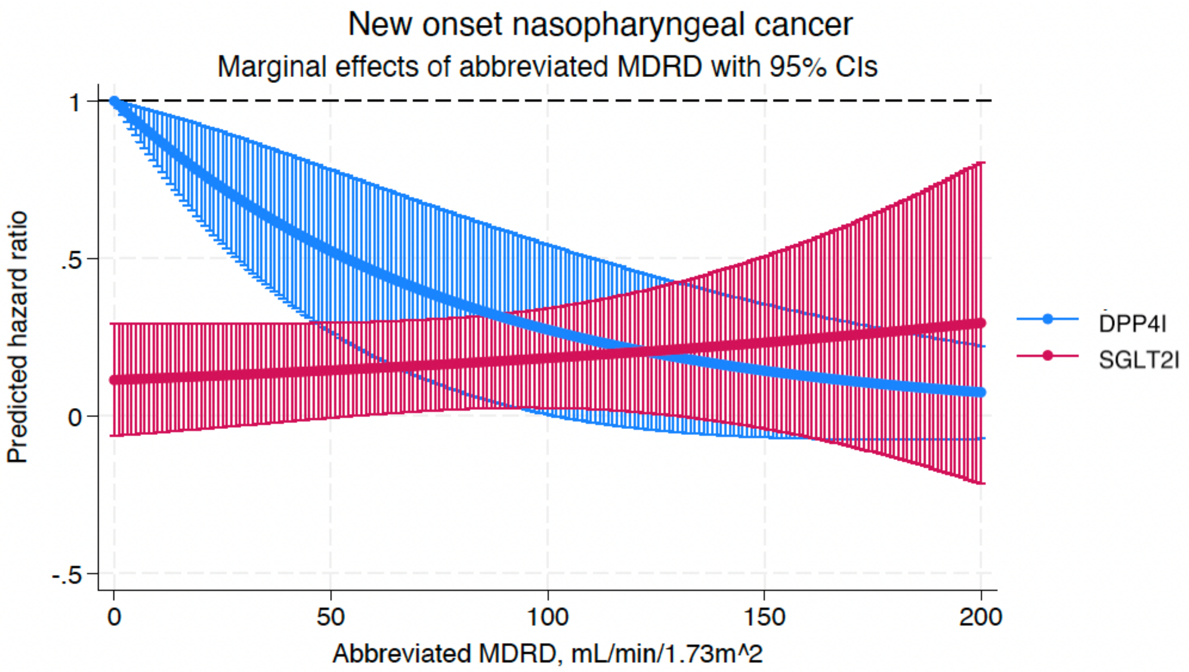

Note: CI: confidence interval.

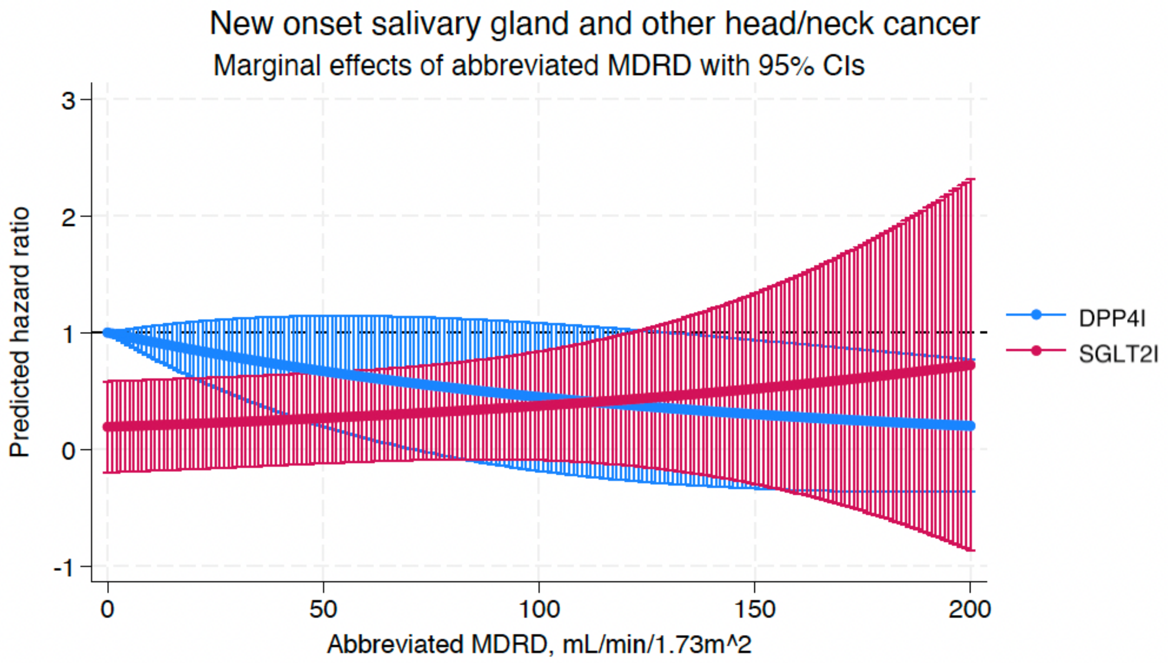

Note: CI: confidence interval.

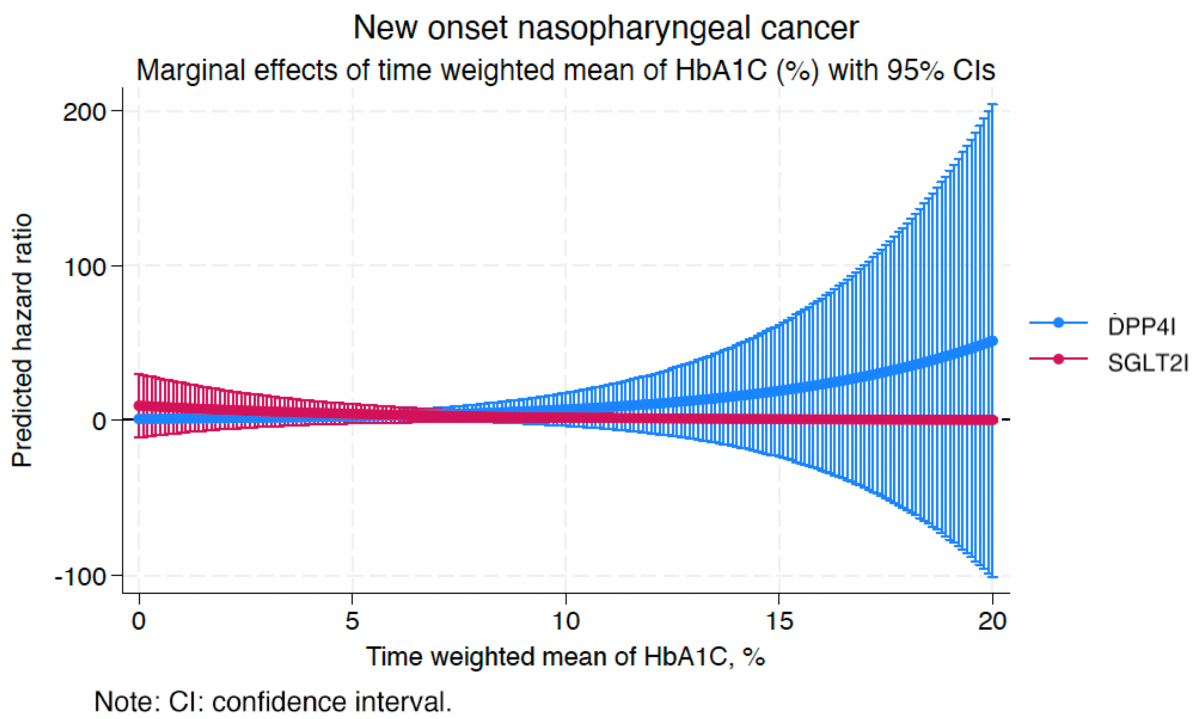

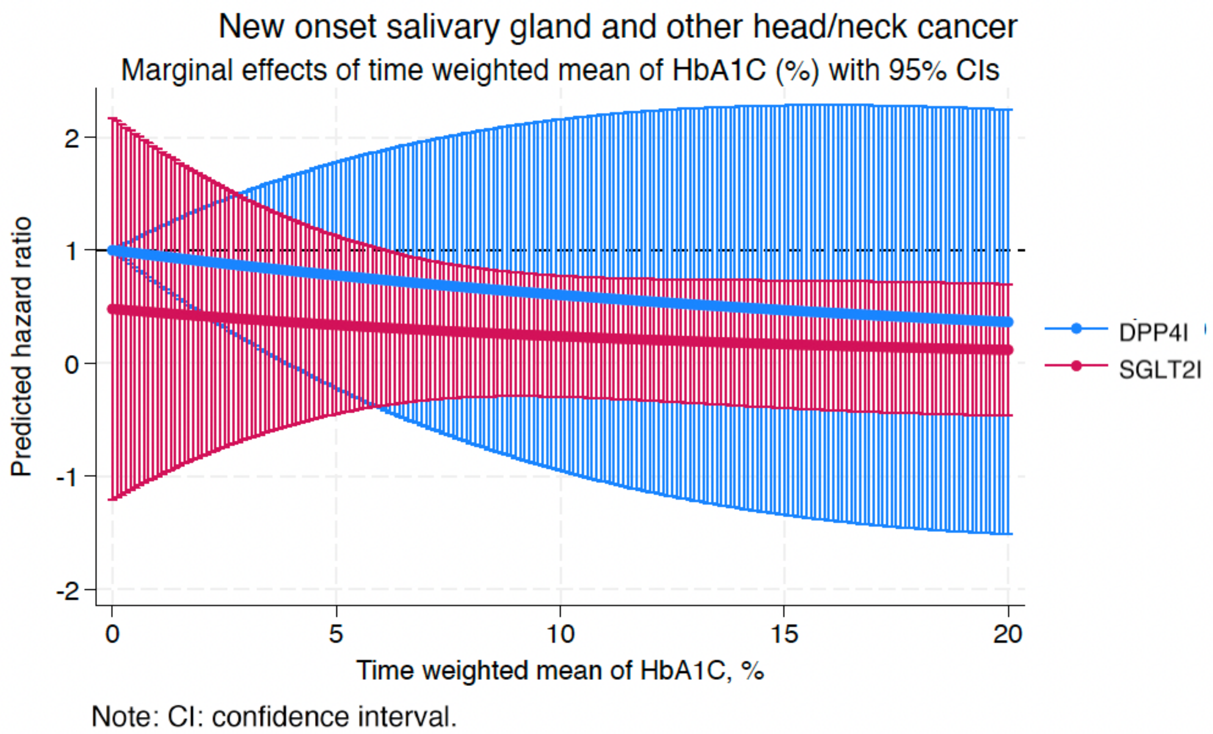

**Supplementary Figure 2. Marginal effects of the demographics, number of anti-diabetic drugs, renal function, and time-weighted mean of HbA1C with 95% CIs on new onset NPC and H&N cancer stratified by drug use in the matched cohort.**
SGLT2I: Sodium-glucose cotransporter-2 inhibitors; DPP4I: Dipeptidyl peptidase-4 inhibitors.

**Supplementary Table 1. The International Classification of Diseases, Clinical Modification (ICD-9-CM) codes for definitions of past comorbidities and outcomes.**

| Adverse outcome of interest |
|---|
| **Nasopharyngeal carcinoma:** 147 |
| **Other head and neck cancer:** 140-149, 160-161 |
| **Past comorbidities** |
| **Cancer**: 140-239 exclude the adverse outcomes of interests |
| **Hypertension**: 401 401.1 401.9 402 402.01 402.1 402.11 402.9 402.91 403 403.01 403.1 403.11 403.9 403.91 404 404.01 404.02 404.03 404.1 404.11 404.12 404.13 404.9 404.91 404.92 404.93 405 405.01 405.09 405.1 405.11 405.19 405.9 405.91 405.99 437.2 + history of uses of anti-hypertensives |
| **Hyperlipidaemia**: 272.0 272.1 272.2 272.3 272.4 + history of uses of lipid-lowering drugs |
| **Liver disease**: 275.1 275.0 571.0 571.1 571.2 571.3 571.40 571.41 571.42 471.49 571.5 571.6 571.8, 571.9 572.0 572.4 572.1 572.3 572.8 573.0 573.4 573.8 573.9 |
| **Heart failure:** 428 428 428.1 428.2 428.2 428.21 428.22 428.23 428.3 428.3 428.31 428.32 428.33 428.4 428.4 428.41 428.42 428.43 428.9 398.91 402.01 402.11 402.91 404.01 404.03 404.11 404.13 404.91 404.93 |
| **Atrial fibrillation:** 427.31 429.4 |
| **Stroke/transient ischemic attack:** 435 435.1 435.2 435.3 435.8 435.9 433.81 433.91 434 436 437 437.1 433.31 433.01 434.01 434.1 434.11 434.9 434.91 437.2 437.3 437.4 437.5 437.6 437.7 437.8 437.9 430 431 432 432.1 432.9 |
| **Ischemic heart disease:** 410.01 410.02 410.1 410.11 410.12 410.2 410.21 410.22 410.3 410.31 410.32 410.4 410.41 410.42 410.5 410.51 410.52 410.6 410.61 410.62 410.7 410.71 410.72 410.8 410.81 410.82 410.9 410.91 410.92 411 411.1 411.8 411.81 411.89 413 413.1 413.9 414 414.01 414.02 414.03 414.04 414.05 414.06 414.07 414.1 414.11 414.12 414.19 414.2 414.3 414.4 414.8 414.9 410 412 |
| **Acute myocardial infarction:** 410 410.01 410.02 410.1 410.11 410.12 410.2 410.21 410.22 410.3 410.31 410.32 410.4 410.41 410.42 410.5 410.51 410.52 410.6 410.61 410.62 410.7 410.71 410.72 410.8 410.81 410.82 410.9 410.91 410.92 |
| **Renal diseases:** 39.95 54.98 403.01 403.11 403.91 404.02 404.03 404.12 404.13 404.92 404.91 582 582 582.1 582.2 582.4 582.8 582.81 582.89 582.9 583 583 583.1 583.2 583.4 583.6 583.7 584.x 585 585.1 585.2 585.3 585.4 585.5 585.6 585.9 586.x 588 588.1 588.8 588.81 588.89 588.9 + MDRD <60 |
| **Diabetic retinopathy:** 250.5 361 362.01 362.02 362.1 362.53 362.81 362.82 362.83 369 379.23 |
| **Diabetic nephropathy:** 250.40 250.41 250.42 250.43 |
| **Diabetic neuropathy**: 250.6, 337.0, 337.1, 354.0 – 355.9, 356.9, 357.2, 358.1, 536.3, 564.5, 596.54, 713.5, 951.0, 951.1, 951.3 |
| **Chronic obstructive pulmonary disease** 491.0 491,1 491.2 491.8 491.9 492.0 492.8 496 |
| **HPV infection:** 079.4 |
| **Primary immunodeficiency:** 042 199.2 279 284.09 284.1 284.11 284.12 284.19 288.01 288.02 288.03 288.09 288.2 288.4 288.50 288.51 288.59 289.53 289.83 996.8 996.80 996.81 996.82 996.83 996.84 996.85 996.86 996.87 996.88 996.89 |

**Supplementary Table 2. Sensitivity analyses for exposure effects of SGLT2I v.s. DPP4I on new onset nasopharyngeal cancer, new onset salivary gland and other head/neck cancer, cancer related mortality, all-cause mortality in the matched cohort using different models.** * for p≤ 0.05, ** for p ≤ 0.01, *** for p ≤ 0.001; SGLT2I: Sodium-glucose cotransporter-2 inhibitors; DPP4I: Dipeptidyl peptidase-4 inhibitors; HR: hazard ratio; CI: confidence interval; PS: propensity score; IPTW: inverse probability of treatment weighting, SIPTW: stable inverse probability of treatment weighting.

| **Model** | **New onset nasopharyngeal cancer HR [95% CI];P value** | **New onset head and neck HR [95% CI];P value** | **Cancer-related mortality HR [95% CI];P value** | **All-cause mortality HR [95% CI];P value** |
|---|---|---|---|---|
| Cause-specific hazard models | 0.46[0.34, 0.66];<0.0001*** | 0.68[0.32, 1.16];0.1265 | 0.31[0.21, 0.35];<0.0001*** | 0.28[0.21, 0.35];<0.0001*** |
| Sub-distribution hazard models | 0.52[0.35, 0.72];<0.0001*** | 0.72[0.44, 1.22];0.1523 | 0.32[0.19, 0.45];<0.0001*** | 0.25[0.20, 0.39];<0.0001*** |
| PS stratification | 0.42[0.29, 0.62];<0.0001*** | 0.75[0.52, 1.31];0.1745 | 0.37[0.24, 0.54];<0.0001*** | 0.36[0.24, 0.62];<0.0001*** |
| PS with IPTW | 0.43[0.32, 0.71];<0.0001*** | 0.68[0.49, 1.19];0.1921 | 0.41[0.31, 0.66];<0.0001*** | 0.44[0.31, 0.59];<0.0001*** |
| PS with SIPTW | 0.66[0.41, 0.79];<0.0001*** | 0.72[0.56, 1.28];0.2128 | 0.42[0.34, 0.72];<0.0001*** | 0.49[0.25, 0.67];<0.0001*** |

**Supplementary Table 3. Sensitivity analysis: Three-arm (only SGLT2I, only DPP4I, and only GLP1a) analysis results using stabilized IPTW, new onset nasopharyngeal cancer, other head & neck cancer, cancer related mortality in the matched cohort (1:1).**
* for p≤ 0.05, ** for p ≤ 0.01, *** for p ≤ 0.001; HR: hazard ratio; CI: confidence interval; SGLT2I: sodium glucose cotransporter-2 inhibitor; DPP4I: dipeptidyl peptidase-4 inhibitor; glucagon-like peptide-1 receptor agonist (GLP1a).

| **New onset nasopharyngeal cancer** | **HR [95% CI]** | **P value** |
|---|---|---|
| SGLT2I vs DPP4I | 1.65 [1.15-2.23] | 0.0005*** |
| SGLT2I vs GLP1a | 1.24 [0.79-1.66] | 0.3451 |
| **New onset head and neck cancer** | | |
| SGLT2I vs DPP4I | 1.01 [0.79-1.12] | 0.4571 |
| SGLT2I vs GLP1a | 1.00 [0.55-1.02] | 0.6823 |
| **Cancer related mortality** | | |
| SGLT2I vs DPP4I | 2.02 [1.68-3.11] | <0.0001*** |
| SGLT2I vs GLP1a | 1.09 [0.56-1.95] | 0.8019 |

**Supplementary Table 4. Sensitivity analysis: Risk of the adverse outcome upon patients with CKD stage 4/5 (eGFR <30), peritoneal dialysis or haemodialysis, excluding patient with prior VT/VF/SCD, excluding patients die within 30 days after drug uses, including patients who were drug abuser, and excluding patients with financial aids, and patients with extreme 10% propensity score at the 2 end.**

* for p≤ 0.05, ** for p ≤ 0.01, *** for p ≤ 0.001; SGLT2I: Sodium-glucose cotransporter-2 inhibitors; DPP4I: Dipeptidyl peptidase-4 inhibitors; HR: hazard ratio; CI: confidence interval.

| SGLT2I v.s. DPP4I | Nasopharyngeal cancer HR[95% CI];P value | Head and neck cancer HR[95% CI];P value | Cancer-related mortality HR[95% CI];P value |
|---|---|---|---|
| 6-months lag time | 0.41[0.27-0.63];<0.0001*** | 0.65[0.37-1.15];0.1397 | 0.24[0.21-0.29];<0.0001*** |
| 1-year lag time | 0.41[0.27-0.63];<0.0001*** | 0.65[0.37-1.15];0.1397 | 0.24[0.21-0.29];<0.0001*** |
| 2-year lag time | 0.41[0.27-0.63];<0.0001*** | 0.67[0.38-1.19];0.1763 | 0.25[0.21-0.29];<0.0001*** |
| Exclude patients with extreme 10% propensity score at the 2 end | 0.41[0.27-0.63];<0.0001*** | 0.65[0.37-1.15];0.1397 | 0.24[0.21-0.29];<0.0001*** |
| Excluding patients with CKD stage 4/5 (eGFR <30), peritoneal dialysis or haemodialysis | 0.53[0.34-0.82];0.0042** | 0.68[0.38-1.20];0.1842 | 0.41[0.34-0.50];<0.0001*** |
| Excluded patients with primary immunodeficiency | 0.40[0.26-0.61];<0.0001*** | 0.65[0.37-1.15];0.1399 | 0.24[0.21-0.29];<0.0001*** |

**Supplementary Table 5. Falsification analysis: Exposure effects of SGLT2I v.s. DPP4I on new onset lower limb fracture (ICD-9 codes: 820-829) in the matched cohort after 1:1 propensity score matching.**

* for p≤ 0.05, ** for p ≤ 0.01, *** for p ≤ 0.001; SGLT2I: Sodium-glucose cotransporter-2 inhibitors; DPP4I: Dipeptidyl peptidase-4 inhibitors; HR: hazard ratio; CI: confidence interval

Adjusted for demographics, past comorbidities, duration of diabetes mellitus, and number of hospitalizations, number of anti-diabetic drugs, non-SGLT2I/DPP4I medications, abbreviated MDRD, HbA1c, fasting glucose.

| | **Number of events, Count(%)** | **Time to lower limb fracture, days (IQR)** | **Lower limb fracture HR[95% CI];P value** |
|---|---|---|---|
| All (N= 57608) | 1061(1.84%) | 1976.4(272.7) | NA |
| SGLT2I (N=28804) | 655(2.27%) | 2008.0(216.6) | 1.29[0.95-1.79];0.6845 |
| DPP4I (N=28804) | 406(1.41%) | 1944.7(315.9) | 1 [Reference] |
